# Spatial-temporal dynamics and recurrence of chikungunya virus in Brazil

**DOI:** 10.1101/2022.08.03.22278339

**Authors:** William M. de Souza, Shirlene T. S. Lima, Leda M. Simões Mello, Darlan S. Candido, Lewis Buss, Charles Whittaker, Ingra M. Claro, Nilani Chandradeva, Fabiana Granja, Ronaldo de Jesus, Poliana S. Lemos, Daniel A. Toledo-Teixeira, Priscilla P. Barbosa, Antonio Carlos L. Firmino, Mariene R. Amorim, Larissa M. F. Duarte, Ivan B. Pessoa, Julia Forato, Irihane L. Vasconcelos, Ana Carolina B. M. Maximo, Emerson L. L. Araújo, Liana Perdigão Mello, Ester C. Sabino, José Luiz Proença-Módena, Nuno R. Faria, Scott C. Weaver.

## Abstract

Chikungunya virus (CHIKV) is an *Aedes* mosquito-borne virus that has caused explosive epidemics linked to acute, chronic, and severe clinical outcomes. Since 2014, Brazil has had the highest number of chikungunya fever (CHIKF) cases in the Americas. Here, we report and contextualize the spatiotemporal dynamic of CHIKF in Brazil and combine genomic, epidemiological, and vector analyses to investigate CHIKF recurrence in several Brazilian states. From 2013 to 2022, CHIKV caused seven epidemic waves across Brazil, affecting 59.5% (3,316 of 5,570) of the country’s municipalities. To date, Ceará State in the northeast has been the most affected, with 81,274 cases during the two largest epidemic waves in 2016 and 2017, and the ongoing third wave in 2022. The 2022 CHIKF recurrence was associated with a new introduction of an East/Central/South African strain. Also, the CHIKV recurrences in Ceará, Tocantins, and Pernambuco States were limited to municipalities with few or no prior reported cases in the previous epidemic waves, suggesting that spatial heterogeneity of CHIKV spread and population immunity may explain the recurrence pattern in the country. In addition, the population density metrics of main CHIKV vector in Brazil, *Ae. aegypti*, were not correlated spatially with locations of CHIKF recurrence in Ceará and Tocantins States. Also, we show that CHIKF disproportionally affected females, and we estimated the case-fatality ratio in Ceará at ∼1.3 deaths per 1,000 cases. These findings more comprehensively describe CHIKV epidemics in Brazil and contribute to understanding CHIKF recurrence in urban settings. Overall, this information may help guide public health policy to mitigate and reduce the burden of urban arboviruses.

## INTRODUCTION

Chikungunya virus (CHIKV) is a major threat to global public health, and it is mainly transmitted among humans by *Aedes aegypti* and *Ae. albopictus* mosquitoes (1). During the past 20 years, CHIKV has caused over 10 million reported cases in more than 125 countries or territories (2). Chikungunya fever (CHIKF) is characterized by acute and chronic human signs and symptoms, typically with severe, often arthralgia but also including some neurological complications and fatal outcomes (2-4). It has been estimated that 1.3 billion people live in areas at-risk for CHIKV transmission (5). Modeling suggests that many more parts of the world might become suitable for CHIKV transmission due to climate change increasing the distribution of *Ae. aegypti* (6). Neither licensed vaccines nor antiviral drugs are available to prevent CHIKV infection or to treat CHIKF.

CHIKV is currently classified into West African, East-Central-South African (ECSA), and Asian genotypes (7). The ECSA genotype recently gave rise to the Indian Ocean lineage (IOL), responsible for 2005-present epidemics in the Indian Ocean Islands, South and Southeast Asia, and Europe (8). The expansion of the ECSA-IOL epidemic has been partly attributed to adaptive mutations in the E1 and E2 envelope glycoproteins, which facilitated adaptation for *Ae. albopictus* infection and transmission (9, 10). CHIKV infection appears to promote life-long immunity, where neutralizing antibodies prevent disease and, to some extent, probably reinfection (11).

CHIKF outbreak recurrences in Africa and Asia are often preceded by long periods of several years to decades with minimal to absent cases. Recurrence can be explained by several factors, including the absence of neutralizing antibodies in younger age groups after periods of epidemiological silence (12, 13). In addition, recurrences of CHIKV in West Africa have been attributed to enzootic CHIKV periodically spilling over into humans entering or living near forests to cause individual cases and small outbreaks (13). The periodicity of these spillover cases appears to be driven by changes in herd immunity among nonhuman primate enzootic hosts (14). However, the dynamics and drivers associated with CHIKV recurrence in urban environments remain poorly understood.

In Brazil, where *Ae. aegypti* and *Ae. albopictus* are widely distributed (15), autochthonous CHIKF cases caused by the Asian and ECSA genotypes were first detected in December 2013 and September 2014, respectively (16). Subsequent genomic investigations indicated the predominance of the ECSA genotype across Brazil’s five geographic regions (17-21). Currently, Brazil has the highest number of CHIKF cases in the Americas (22). Here, we contextualize the CHIKV spread in Brazil from 2013 to 2022 and combine epidemiological, genomic, and vector density population analysis to describe and investigate CHIKV recurrence.

## RESULTS

### Spatio-temporal dynamics of chikungunya virus across Brazil

Up to 4 June 2022, 253,545 laboratory-confirmed CHIKF cases were reported to the Brazilian Ministry of Health across 3,316 out of 5,570 municipalities (59.5%) in all 26 states and the Federal District. CHIKF cases were mainly distributed across seven large epidemic waves, which resulted in 24,097 to 44,604 confirmed cases annually (**Fig. 1a** and **Table S1**). The epidemic peak of cases in most affected Brazilian states varied annually between February and July (**Table S1**). Interestingly, we found that Google searches of the search term “chikungunya” in Brazil captured these seven large epidemic waves, with a high correlation (ρ=0.74, Spearman’s rank correlation coefficient) between Google Trends activity and the number of CHIKF cases in the states most affected (**Fig S1**). Since early May 2022 (epidemiological week [EW] 18), we observed a decrease of 75.1% in CHIKF cases reported (EW-18 = 2,493 cases and EW-22 = 620 cases), likely due to a shortage of diagnostic kits for CHIKV, dengue virus (DENV), and Zika virus (ZIKV) in Brazil (23) (**Fig. 1a**). Northeast Brazil was the region most affected by CHIKF, with 63.5% (n = 160,909) of all reported cases between 2013 to 2022 (**Fig. 1a**). Ceará State had the highest number of cases (n = 45,417) and an accumulated incidence of 501.4 cases per 100,000 inhabitants from 2013 to 2022 (**Fig. 1a-b**).

**Fig. 1.**
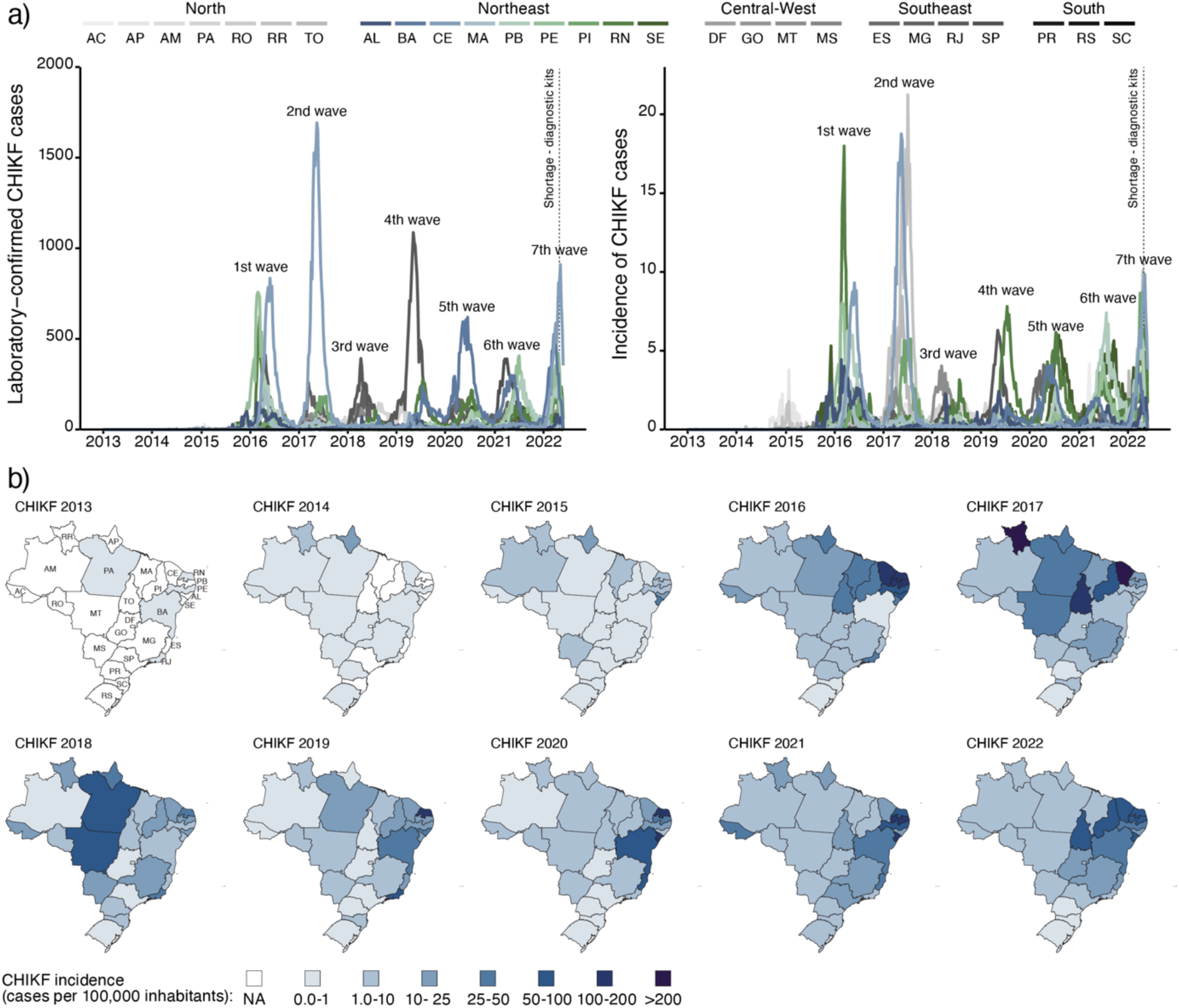
Spatial-temporal dynamics of chikungunya virus between 2013 and 2022 in Brazil. **a)** Number of laboratory-confirmed CHIKF cases (left) and incidence based on laboratory-confirmed CHIKF cases (right) per epidemiological week in all 26 Brazilian States and the Federal District from epidemiological week 1 of 2013 (30 December 2012 to 5 January 2013) to epidemiological week 22 of 2022 (29 May 2022 to 4 June 2022). **b)** Maps colored according to the incidence of laboratory-confirmed CHIKF cases per state reported to the Brazilian Ministry of Health. Map for 2022 is limited to epidemiological weeks 1-22. AC, Acre; AL, Alagoas; AM, Amazonas; AP, Amapá; BA, Bahia; CE, Ceará; DF, Distrito Federal; ES, Espírito Santo; GO, Goiás; MA, Maranhão; MG, Minas Gerais; MS, Mato Grosso do Sul; MT, Mato Grosso; PA, Pará; PB, Paraíba; PE, Pernambuco; PI, Piauí; PR, Paraná; RJ, Rio de Janeiro; RN, Rio Grande do Norte; RO, Rondônia; RR, Roraima; RS, Rio Grande do Sul; SC, Santa Catarina; SE, Sergipe; SP, São Paulo; TO, Tocantins.

### Recurrence of chikungunya fever in Ceará State after two large epidemic waves

To evaluate the epidemiological dynamics in Ceará State, we analyzed the individualized data of all patients tested for CHIKV (n = 146,887) in the Central Laboratory of Public Health up to 31 May 2022. Ceará State experienced three CHIKF waves since the first autochthonous case was reported on 6 March 2015 in Fortaleza, the state’s capital and most populous city (**Fig. 2a**). The first and second waves in Ceará State had the epidemic peak (> 30% of cases) in May and caused 17,012 and 40,596 cases, respectively. After four years (December 2017 to December 2021) of CHIKF incidence ≤1 case per 100,000 inhabitants, the disease reoccurred in a third epidemic, causing 19,810 cases. Comparing the first five months during the years of CHIKV transmission in Ceará State, the ongoing third wave caused 2.2 times more cases than the first wave, but 1.6 times fewer cases than the second wave (**Fig. 2b**). In addition, our analysis of the age-sex structure of CHIKF cases in the three epidemic waves revealed a significantly higher combined incidence (2.2 to 3 times) in adult female age groups (20 to 59 years old) compared to males (**Fig. 2c**). Based on analysis of a set of real-time RT-PCR-positive cases for the ongoing third wave (n = 638 patients), no differences were observed in mean cycle threshold (Ct) values between sexes (**Fig. S2**). In addition, we found that young patients had significantly lower median Ct values, indicating that many young patients have more viral nucleic acid in serum compared to adults and the elderly (**Fig. S2**).

**Fig. 2.**
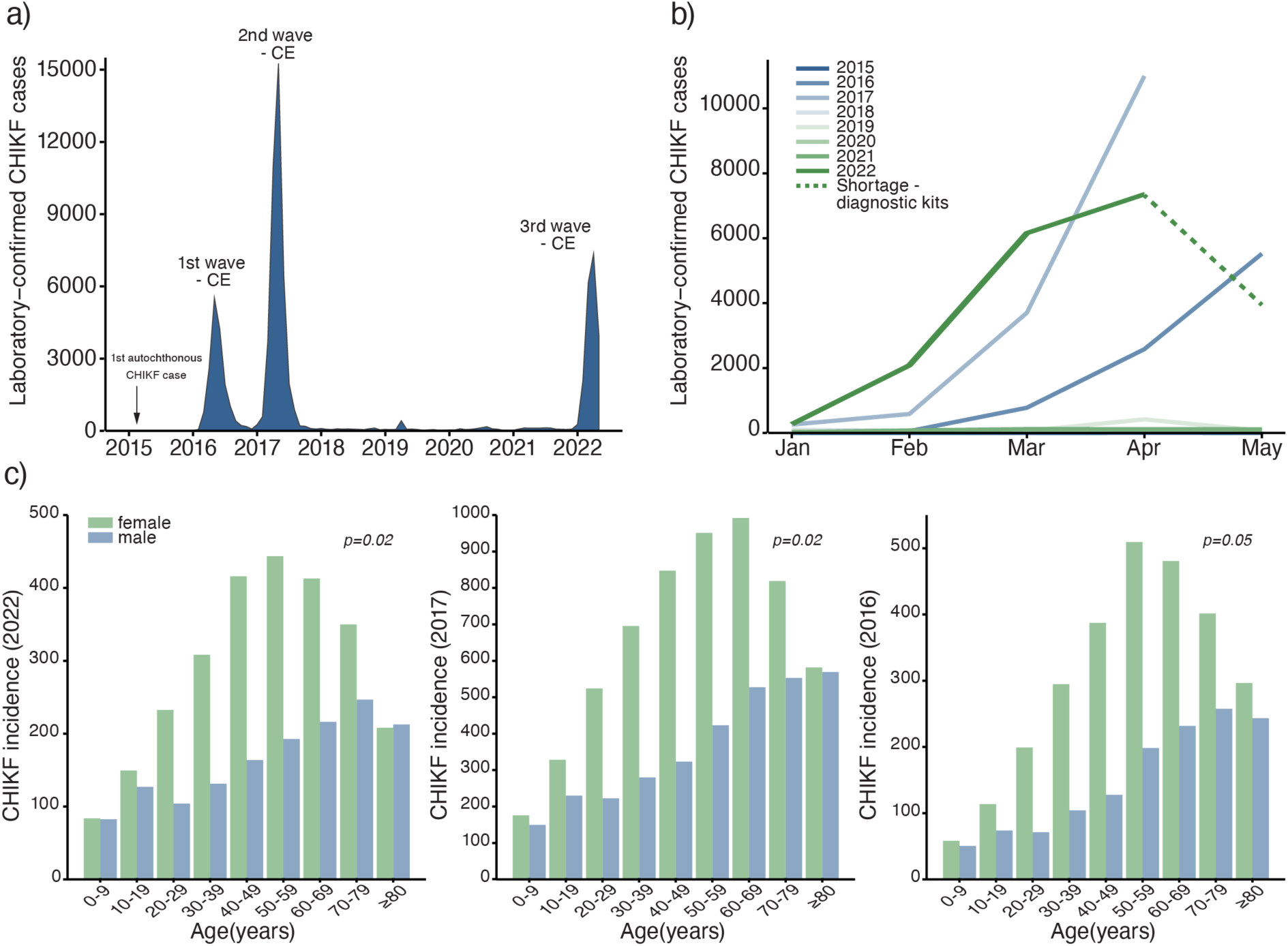
Chikungunya virus waves in Ceará state, Brazil. **a)** Number of laboratory-confirmed CHIKF cases per month from 6 March 2015 to 31 May 2022. **b)** Number of laboratory-confirmed CHIKF cases per year (2015-2022) from 1 January to 31 May. **c)** CHIKF incidence based on age-sex distribution of epidemic waves in 2022 (left), 2017 (center), and 2016 (right). The statistical difference between sexes in all age groups was calculated by two-way ANOVA and post-hoc test were performed correcting for multiple comparisons using the Dunn–Bonferroni method. The normality assumption was checked based on the Shapiro-Wilk Test. The significance level was determined as p<0.05.

### Recurrence of chikungunya virus in Brazil was associated with viral transmission in regions and municipalities less affected in previous waves

To investigate which municipalities were most affected during the three CHIKV waves in Ceará state, we analyzed the spatial distribution and incidence of CHIKF cases across 184 municipalities in all seven mesoregions from 2015 to 2022. We observed that CHIKF recurrence in 2022 has predominantly affected municipalities located in the south of Ceará State (**Fig. 3a**). The third wave is occurring in a small number of municipalities (n = 37) with an incidence of ≥100 CHIKF cases per 100,000 inhabitants compared to 100 municipalities that reported ≥100 CHIKF cases per 100,000 inhabitants in the first and second waves. In addition, five out of seven Ceará mesoregions reported a total of >800 cases and an incidence of >100 cases per 100,000 population per month during the first and/or second CHIKF waves, except *Centro-Sul Cearense* and *Sul Cearense*; the latter is currently the most affected mesoregion in 2022 (**Fig. 3b**). Comparing CHIKF incidence caused by the first and second waves (2016 and 2017) against the third wave (2022), we found that municipalities most affected during the recurrence were less affected during the first two epidemics. Also, we found a similar pattern in Pernambuco and Tocantins States, which were affected by CHIKF recurrence in 2021 and 2022, respectively (**Fig. 3c**). These data show that CHIKF recurrence occurred primarily in regions and municipalities that had been less affected during previous epidemic waves in Brazil.

**Fig. 3.**
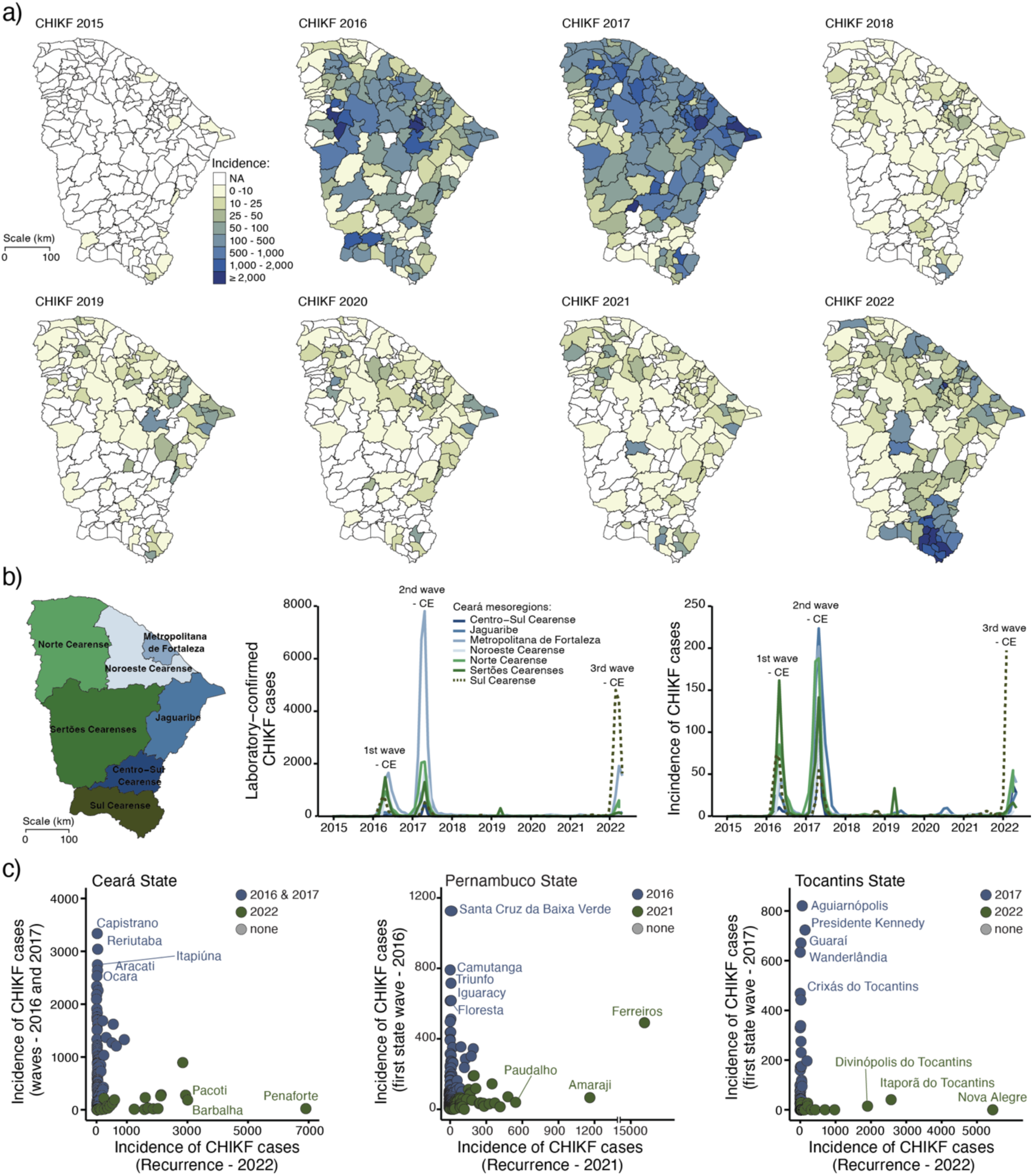
Spatio-temporal distribution of recurrence of chikungunya virus. **a)** Spatio-temporal distribution of annual incidence based on laboratory-confirmed CHIKF cases per municipality (n = 184 municipalities) of Ceará State from 2015 to 2022. CHIKF incidence in Ceará State in 2022 includes data only until 31 May. **b)** Map of mesoregions of Ceará State, Brazil (left). Number of CHIKF cases (center) and CHIKF incidence (right) per mesoregions of Ceará State from 1 January 2015 to 31 May 2022. **c)** Comparison of CHIKF incidence caused by the previous CHIKV waves and recurrence in Ceará (left), Tocantins (center), and Pernambuco States (right).

### Chikungunya virus can cause explosive outbreaks with a higher number of cases and deaths compared to dengue virus

Since 2016, CHIKV has caused 103 confirmed-laboratory deaths across 28 municipalities in Ceará State through 31 May 2022, with 53.4% of deaths reported in Fortaleza city (**Fig. 4a**). The peaks of deaths overlapped with the peaks in the number of cases during the three CHIKF waves (**Fig. 4b**). The cumulative case fatality ratio (CFR) for 2016 to 2022 was ∼1.3 deaths per 1,000 cases. In addition, we found a positive correlation between deaths and cases per month (**Fig. 4c**). We found that CHIKF deaths occurred predominantly in females (male-to-female ratio = 0.78), but no statistical differences were observed between sexes. Also, we identified a U-shaped pattern of age-associated mortality (**Fig. 4d**). In comparison, DENV caused 72 deaths in 29 municipalities of Ceará State during the same period, and most cases (46.8%) were also reported in Fortaleza city. No correlation was observed between DENV laboratory-confirmed deaths and monthly cases (**Fig. S3**). The CFR for DENV was estimated at ∼1.1 deaths per 1,000 laboratory-confirmed cases. DENV deaths affected more males (male-to-female ratio = 1.1), especially older individuals, but without a statistical difference between sexes (**Fig. S3**). No significant difference was observed between the CFRs of CHIKV and DENV.

**Fig. 4.**
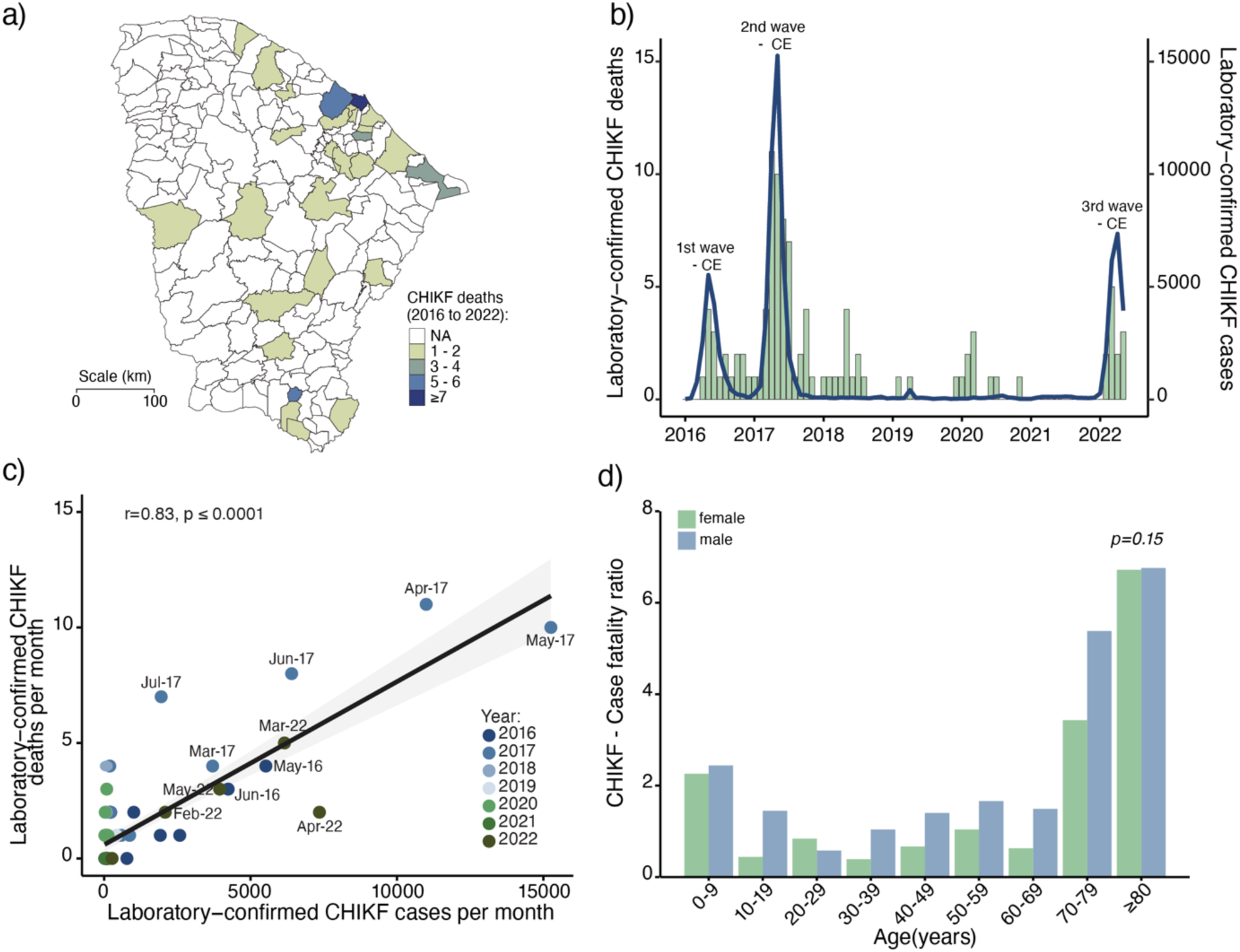
Chikungunya deaths in Ceará State, Brazil. **a)** Spatial distribution of laboratory-confirmed CHIKF deaths per municipality in Ceará State from 2016 to 2022. **b)** Number of laboratory-confirmed CHIKF deaths (blue line) and cases (green bars) per month from 1 January 2016 to 31 May 2022. **b)** The Pearson’s correlation coefficients of laboratory-confirmed CHIKF deaths per month per laboratory-confirmed CHIKF cases per month from 2016 to 2022 in Ceará State. **d)** The statistical difference between sexes in all age groups was calculated by two-way ANOVA and a post-hoc test was performed correcting for multiple comparisons using the Dunn–Bonferroni method. The significance level was determined as p<0.05.

### A distinct lineage of the CHIKV-ECSA genotype was associated with the chikungunya virus recurrence in Ceará State

We next investigated the genetic diversity of CHIKV in Ceará State in 2022. We sequenced 61 genomes from the recent outbreak collected from February to March 2022, including cases from the three mesoregions: *Metropolitana de Fortaleza* (n = 3), *Noroeste Cearense* (n = 6), and *Sul Cearense* (n = 53) (**Fig. 5a** and **Table Supplementary 1**). All genomes had ≥ 85% coverage and a mean depth of coverage of at least 20×. A regression of genetic divergence from root to tip against sampling dates confirmed the strong temporal signal of our genomic dataset (**Fig. 5b**). Next, we estimated a timescale for the evolution of the CHIKV clade in 2022 using a best-fitting molecular clock model. Phylogenetic analysis revealed that an introduction of a new CHIKV-ECSA lineage strain caused the third epidemic wave in Ceará State (**Fig. 5a**). This is most closely related to CHIKV strains circulating recently in São Paulo State, and the most recent common ancestor of the CHIKV-ECSA lineage associated with recurrence in Ceará State was estimated on or around 17 July 2021 (95% Bayesian credible interval = 15 October 2020 to 14 October 2021). We did not find previously described mutations associated with enhanced transmission potential for *Ae. albopictus* mosquitoes (e.g., E1-A226V and E1-T98A) in the CHIKV strains circulating in 2022, consistent with a lack of incrimination of this species in transmission in the Americas. Also, we did not find epistatic interactions that control A226V penetrance (i.e., E1-211 and E1-98), which are associated with adaptation to a new vector, and thus can restrict epidemic emergence (24).

**Fig. 5.**
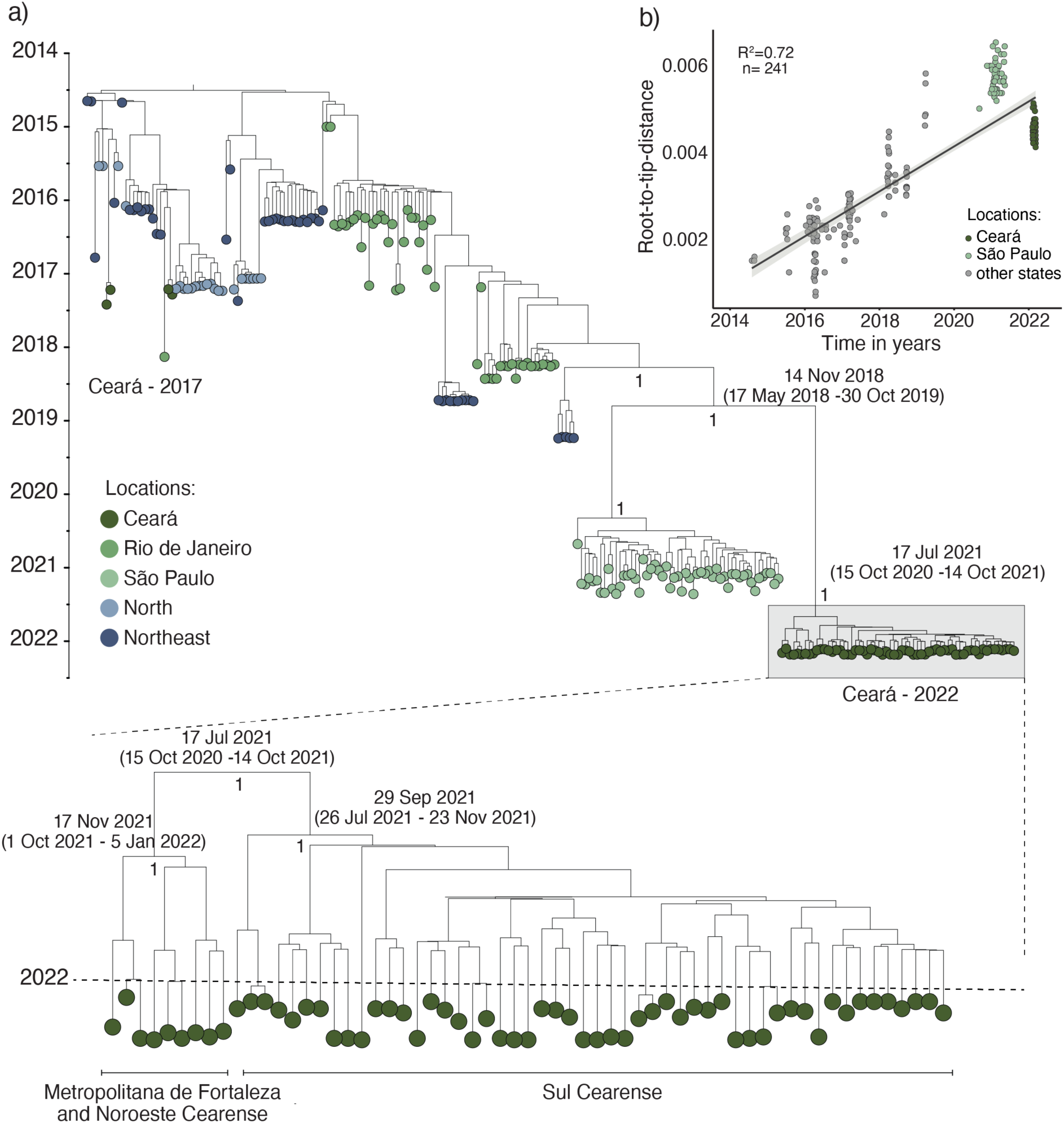
Phylogenetic analysis of East-Central-South African genotype of chikungunya virus in Brazil. **a)** Maximum clade credibility tree includes 241 CHIKV genomes from the ECSA genotype, including 61 new CHIKV genomes from Ceará State generated in this study. Tips are colored according to the source region or state of each sample. A strict molecular clock approach was used for generating the time-rooted tree. **b)** Regression of sequence sampling dates against root-to-tip genetic distances in a maximum likelihood phylogeny of the CHIKV ECSA genotype in Brazil. Sequences are colored according to the three locations (Ceará, São Paulo, and other Brazilian states). Posterior probability scores are shown next to key well-supported nodes.

### Vector density population was not associated with recurrence of chikungunya virus in Ceará and Tocantins States

*Aedes aegypti* is the main vector of CHIKV and DENV in Brazil (25). An increase in vector population density has been associated with the rise of dengue incidence (26, 27); thus, we sought to determine whether a recent increase in *Ae. aegypti* population density could correlate with the increase in CHIKV transmission. For this, we evaluated the Breteau (BI) and House indices (HI) for all Ceará State municipalities and 83.5% (116 of 139) of Tocantins State municipalities from 3 January to 21 February 2022. In Ceará State, we found that 39.1% (72 of 184) of municipalities presented a BI and/or HI above 1, which triggers a risk alert for *Ae. aegypti* -borne virus outbreaks according to the National Dengue Control Programme in Brazil (28) (**Fig. 6a**). In contrast, we identified that 85.3% (99 of 116) of Tocantins State municipalities presented a BI and/or HI above 1 (**Fig. 6b**). However, no correlation was observed between *Ae. aegypti* indices (HI and BI) and CHIKF incidence in Ceará and Tocantins municipalities in 2022 (**Fig.6b**). Also, we did not find a correlation between *Ae. aegypti* indices (HI and BI) and dengue in Ceará State in 2022 (**Fig.S4**). We did not evaluate the correlation between CHIKF recurrence and *Ae. aegypti* indices in Pernambuco due to the absence of vector data for the recurrence period in 2021. These data suggest that vector density, as measured by traditional indices, was not a major driver of the recurrence of CHIKF in Ceará and Tocantins States.

**Fig. 6.**
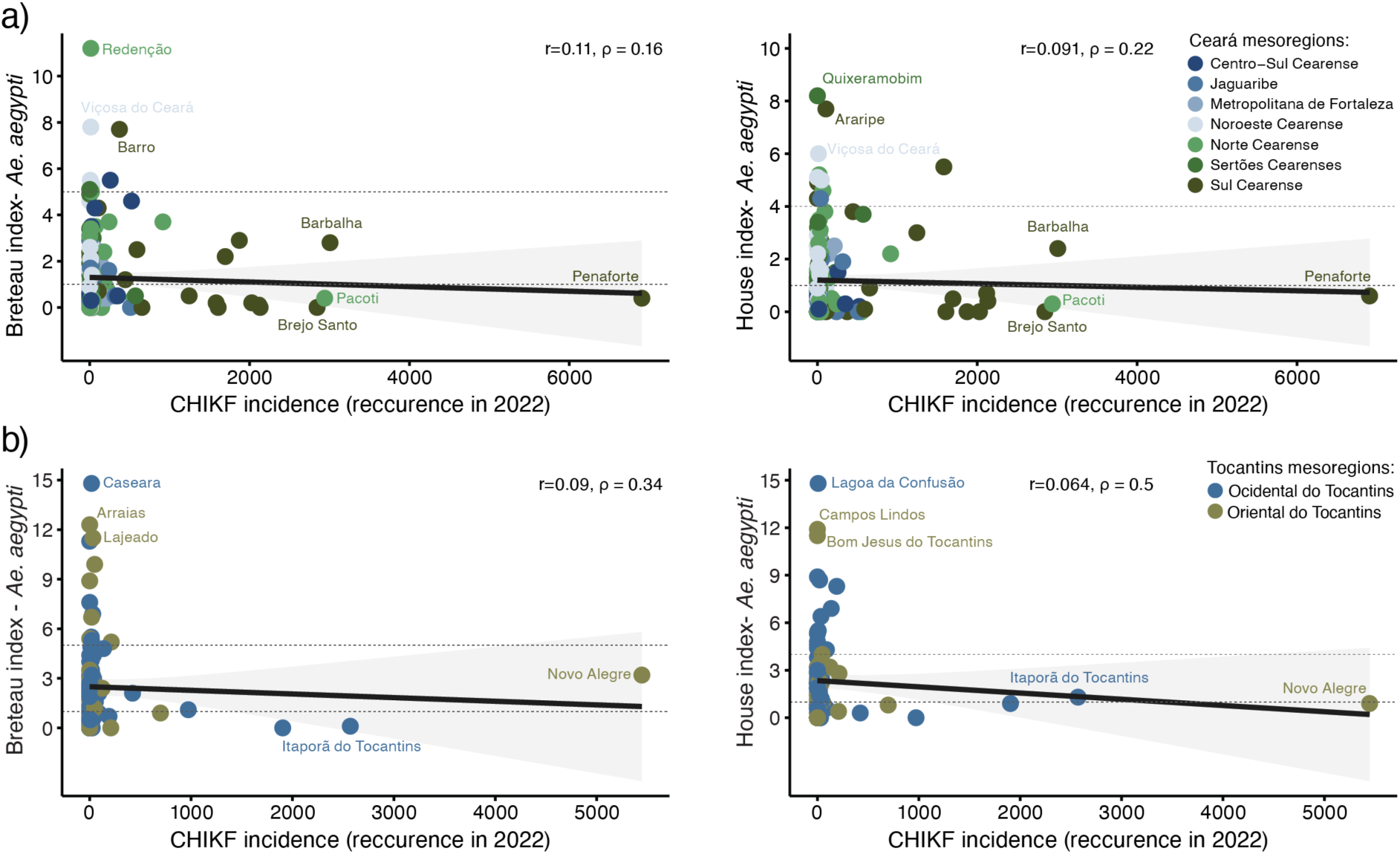
Correlation between *Ae. aegypti* population density indices and chikungunya incidence in Ceará and Tocantins States in 2022. Correlation coefficients of Breteau index (left) and House index (right) per CHIKF incidence in 2022 in Ceará State municipalities (**a**) and Tocantins State municipalities (**b**). The correlation was calculated using Spearman’s rank correlation coefficient. The circles were colored based on mesoregions. The upper dash line indicates the risk situation (Breteau index = 5 and House index = 4), and the bottom dash line denotes the alert situation (Breteau index and House index > 1) according to the National Dengue Control Programme in Brazil (28).

## DISCUSSION

We describe and contextualize the spatial and temporal dynamics of seven large CHIKF epidemic waves in Brazil between 2013 and 2022, and investigate the recurrence after a four-year hiatus in three Brazilian states. We found that Ceará was the most affected Brazilian state, where the recurrence in 2022 resulted from a new lineage of CHIKV-ECSA introduced around mid-2021 that was distinct to previous lineages circulating in this state (3). However, this new viral lineage does not alone explain the observed recurrence because previous CHIKV infection produces a robust humoral response that prevents reinfection, even against other genotypes (11, 29). Instead, our analysis suggests that the spatial heterogeneity of CHIKV spread during the first waves can help explain recent CHIKF recent recurrence. We found that municipalities in Ceará, Pernambuco, and Tocantins States that experienced CHIKF recurrence were affected less or completely unaffected by previous waves. In contrast, regions deeply affected in previous waves had no higher incidence during the 2022 CHIKF recurrence. Therefore, it is plausible that the populations in municipalities most affected by CHIKF waves presented some level of immune protection against disease and/or transmission that temporarily prevented the re-occurrence of explosive CHIKF outbreaks. In contrast, the populations from municipalities less exposed to earlier CHIKV waves, remained more susceptible.

Our results show that CHIKF affected females disproportionally, consistent with previous findings that suggest females are 1.5 times more likely to become infected than males (30). Here, we also estimated the case-fatality ratio of CHIKF in Ceará at ∼ 1.3 deaths per 1,000 cases, which is similar to that previously described in Réunion Island in the Indian Ocean in 2005–2006 (1 death per 1,000 cases) with a higher sensitivity surveillance system (31), which was also consistent with iterative external studies and serosurveys (32). Collectively, our results indicate that CHIKF outbreaks can be explosive and result in a large number of cases and a case-fatality ratio similar to that seen in DENV epidemics. Therefore, mechanisms and potential biomarkers associated with severe CHIKV outcomes need to be investigated to support the development of antiviral therapies, improve clinical management, and reduce the CHIKF burden.

We found that peaks of CHIKF cases in the epidemic waves occur mainly between February and June in Brazil, coinciding with the rainy season and higher temperatures. These wet and warm periods have been described as critical drivers of the magnitude and seasonality of DENV transmission based on affecting the mosquito’s reproduction, survival, biting rates, and the adult vector population density (33-35). Consequently, this can increase the risk and the dynamic of arbovirus transmission, such as CHIKV and DENV. Also, in our analysis, *Ae. aegypti* population density metrics were not correlated spatially with locations of CHIKF recurrence in Ceará and Tocantins States. These findings underscore that vector population density thresholds (e.g., HI and BI) need to be improved by incorporating herd immunity data to better predict outbreaks transmitted by *Ae. aegypti* and *Ae. albopictus* (36). More robust modeling that accounts for mosquito population dynamics, climate data, prior immunity to CHIKV, and the distribution of *Ae. aegypti* control interventions across the country could help to quantify CHIKF burden and evaluate the impact of vector density on CHIKF recurrence.

There are several limitations to our study. First, the lack of CHIKV genomic sequences from other Brazilian states prevents us from drawing accurate conclusions on the date of introduction and geographic origins of the new lineage in Ceará State. Here, we propose that the implementation of screening in blood donors combined with CHIKV genome sequencing (37) could provide unique data to understand the epidemiological and evolutionary dynamics of CHIKV in Brazil. Second, the absence of CHIKV seroprevalence studies at the state and national levels in Brazil limits estimates of the susceptibility and herd immunity in the population. For example, some CHIKV-endemic countries, such as India, have an overall prevalence of IgG antibodies of 18% (38). In contrast, our data show that no more than 3% of the population from municipalities deeply affected received a laboratory diagnosis for CHIKF in the first and second waves in Ceará State. Several factors could explain these discrepancies, such as oligo- and asymptomatic cases (up to 50% of cases)(39), challenges of the syndromic surveillance systems due to the co-circulation of ZIKV and DENV which cause similar symptoms to CHIKV (40), and variable healthcare-seeking behaviors. Also, the shortage of diagnostic kits for arboviruses during the epidemic may underestimate the CHIKF burden in 2022.

In conclusion, we have provided a comprehensive assessment of CHIKF epidemics and re-occurrence in Brazil. Our findings provide an important context about the dynamics and drivers of CHIKF and may inform future studies and public health policy focusing on the strategies to mitigate the impact of new epidemic waves in urban settings and immunization programs.

## METHODS

### Definition of cases and ethical statement

Laboratory-confirmed cases of CHIKF or DENV were defined as a patient with one positive laboratory result for CHIKV or DENV either by reverse-transcription quantitative polymerase chain reaction (RT-qPCR), immunoglobulin M (IgM) detection, immunoglobulin G (IgG), viral isolation (e.g., C6/36 cells), and/or non-structural protein 1 (NS1) antigen for DENV. No clinical epidemiological cases were included in this study. This study was approved by the ethics committees from the University of Campinas, Brazil (approval number #53910221.0.0000.5404).

### Epidemiological analysis

National epidemiological data of CHIKF laboratory-confirmed cases were obtained from the Brazilian Ministry of Health. This dataset includes the aggregate number of cases of CHIKF per epidemiological week from all municipalities of Brazil from epidemiological week 10 (3 to 9 March) in 2013 to epidemiological week 22 in 2022 (20 May to 06 June). Ceará State epidemiological data of CHIKF and DENV from 1 January 2015 to 31 May 2022 were obtained from the Laboratory Public Health of Ceará, Brazil. This data includes individualized and de-identified information, such as age, sex, municipality residence, date of symptoms, date of collection, and diagnosis method. Incidences were calculated based on the estimated population of Ceará State from 2015 to 2021, as reported by the Brazilian Institute of Geography and Statistics (www.ibge.gov.br).

### Digital surveillance of chikungunya virus in Brazil

We used the Google Trends tool to compile the monthly fraction of online searches for the term “chikungunya”, which originated from Brazil from 1 January 2013 to 31 May 2022 and plotted it in a time series. In addition, we evaluated the correlation between the Google Trends activity and laboratory-confirmed CHIKF cases per Brazilian federal units using Spearman’s rank correlation test in the R software.

### Sample collection

Sera were collected from patients with samples testing positive for CHIKV by RT-qPCR between 7 February to 9 March 2022 in Ceará State, Brazil. Basic clinical and demographic data were collected through the Brazilian Laboratorial Environment Management System (*Gerenciador de Ambiente Laboratorial*). Anonymized patient information data of all samples used in the current study are provided in **Table S2**.

### A real-time reverse transcription-polymerase chain reaction (real-time RT-PCR) for chikungunya virus

Serum samples were extracted using the Maxwell® RSC Viral Total Nucleic Acid Purification Kit (Promega, USA). RNA extracted were tested by specific real-time RT-PCR for CHIKV as described previously (41). The assays were performed with TaqMan™ Fast Virus 1-Step Master Mix (Applied Biosystems, USA) on the StepOnePlus™ Real-Time PCR System (Applied Biosystems, USA).

### Chikungunya virus genome sequencing and assembly genome

Positive RNA samples by RT-qPCR with cycle threshold (Ct) values <30 were submitted for CHIKV genome sequencing using a targeted multiplex PCR scheme and the MinION platform (Oxford Nanopore Technologies, UK), as described elsewhere (42). PCR products were cleaned using AmpureXP purification beads (Beckman Coulter, United Kingdom) and quantified using fluorimetry with the Qubit dsDNA High Sensitivity assay on the Qubit 3.0 instrument (Life Technologies, USA). Amplicons from each sample were normalized, pooled, and barcoded using the Rapid Barcoding Kit 96 kit (EXP-NBD 196, Oxford Nanopore Technologies, UK). Next, sequencing libraries were generated using the SQK-LSK109 Kit (Oxford Nanopore Technologies, Oxford, UK) and were loaded onto an R9.4.1 flow-cell (Oxford Nanopore Technologies, UK). Then, FAST5 files containing the raw signal data were base-called, demultiplexed, and trimmed using Guppy version 4.4.1 (Oxford Nanopore Technologies, UK). The reads were aligned against the CHIKV strain C302F/2016/BR (GenBank accession no. KY055011) using minimap2 version 2.17.r941 (43) and converted to a sorted BAM file using SAMtools (44). Length filtering, quality testing, and primmer trimming were performed for each barcode using guppyplex. Variants were detected with medaka_variants and the consensus sequence were built using margin_medaka_consensus (Oxford Nanopore Technologies, UK). Genome regions with a depth coverage below 20-fold were represented with “N” characters.

### Collation of chikungunya virus complete genome datasets

A total of 61 new CHIKV sequences with > 85% genome coverage were generated in this study, and appended to 180 other Brazilian CHIKV complete coding sequences and classified into ECSA genotypes available in the GenBank database until 31 May 2022. Then, the multiple sequence alignment was built as MAFFT version 7.450 (45), and manual adjustment was conducted using Geneious Prime 2020.2.3. Subsequently, the dataset was screened for recombination events using all available methods in RDP version 4 (46). No evidence of recombination was found.

### Phylogenetic analysis

Maximum likelihood (ML) phylogeny using IQ-TREE version 2 under a GTR + I + γ model as determined by ModelFinder (47, 48). Statistical support for nodes of the ML phylogeny was assessed using an ultrafast-bootstrap approach with 1,000 replicates. We then regressed root-to-tip genetic divergence against sampling dates to investigate the temporal signal and identify sequences with low data quality of our datasets, such as assembling issues, sample contamination, data annotation errors, sequencing, and alignment errors (49). No obvious outliers were identified in this step. Dated-phylogenetic trees were estimated using BEAST v.1.10.4 under a GTR + I + γ model (50), strict molecular clock model and a Skygrid tree prior (51), and using BEAGLE to enhance computation speed (52). Evolutionary analyses were run independently in duplicate for 50 million steps, sampling parameters, and trees every 10,000 steps. Maximum clade credibility summary trees were generated using TreeAnnotator v.1.10.

### Breteau index and House index for Ceará and Tocantins States

The Breteau index (BI) and House index (HI) for all 184 Ceará State municipalities and 116 out of 139 Tocantins State municipalities were performed between 3 January to 21 February 2022 as part of *Ae. aegypti* Infestation Index rapid Survey (53). BI is a number of water containers containing *Ae. aegypti* larvae per 100 houses, and HI is the percentage of houses with infested containers. These data were obtained from the Brazilian Ministry of Health through the Right to Information Law (Law 12.527-Brazil) under process (#25072.015694/2022-81).

## Data Availability

All statistical computing analyses were conducted using the R project. No custom code was developed. New sequences have been deposited in GenBank with accession numbers (will be made available upon acceptance of the publication).

## Data availability

All statistical computing analyses were conducted using the R project (54). R packages necessary for analysis and visualization include: tidyverse (55), raster(56), tmap(57), ggbreak(58), ggpubr(59), scico(60) and sf(61). No custom code was developed. New sequences have been deposited in GenBank with accession numbers (will be made available upon acceptance of the publication).

## ACKNOWLEDGMENTS

WMS is supported by a Global Virus Network fellowship and Burroughs Wellcome Fund – Climate change and human health seed grants (#1022448). SCW is supported by National Institutes of Health grant AI12094. This project was supported by the Medical Research Council and FAPESP - Brazil-UK Centre for (Arbo)virus Discovery, Diagnosis, Genomics, and Epidemiology partnership award (MR/S0195/1 and FAPESP 2018/14389-0). NC acknowledges funding from the MRC Centre for Global Infectious Disease Analysis (reference MR/R015600/1), jointly funded by the UK Medical Research Council (MRC) and the UK Foreign, Commonwealth & Development Office (FCDO), under the MRC/FCDO Concordat agreement and is also part of the EDCTP2 programme by the European Union. NRF is supported by a Wellcome Trust and Royal Society Sir Henry Dale Fellowship (grant no. 204311/Z/16/Z). JLPM is supported by National Council for Scientific and Technological Development (CNPq), grant no 305628/2020-8. MRA and PPB were supported by Coordination for the Improvement of Higher Education Personnel scholarships (CAPES# 88887.356527/2019-00 and 88887.661921/2022-00). DATT is supported by CNPq scholarship (#141844/2019-1).

## AUTHOR CONTRIBUTIONS

WMS, STSL, NRF and SCW conceptualized the study. WMS, STSL, LMSM, FG, RJ, PSL, DATT, ACLF, MRA, OPB, LMFD, IBP, JF, ILV, ACBM, ELLA, and LPM contributed to the acquisition of data. WMS, DSC, IMC, RJ, and NC contributed to the data analysis. WMS, STSL, LB, DSC, CW, NRF, and SCW contributed to data interpretation. WMS, STSL, LB, DSC, CW, and NRF drafted the manuscript. WMS, LB, CW, NC, NRF, and SCW revised the manuscript. Funding: WMS, ECS, JLPM, NRF, and SCW acquired funding for the study. All authors read and approved the final version of the manuscript and had access to all the data in the study.

## DECLARATION OF INTERESTS

The authors declare no competing interests.

## Notes

### Competing Interest Statement

The authors have declared no competing interest.

### Author Declarations

This study was approved by the ethics committees from the University of Campinas, Brazil (approval number #53910221.0.0000.5404).

