## Supplementary Figures 1-4 and Table Supplementary 1-2 for "Spatial-temporal dynamics and recurrence of chikungunya virus in Brazil"

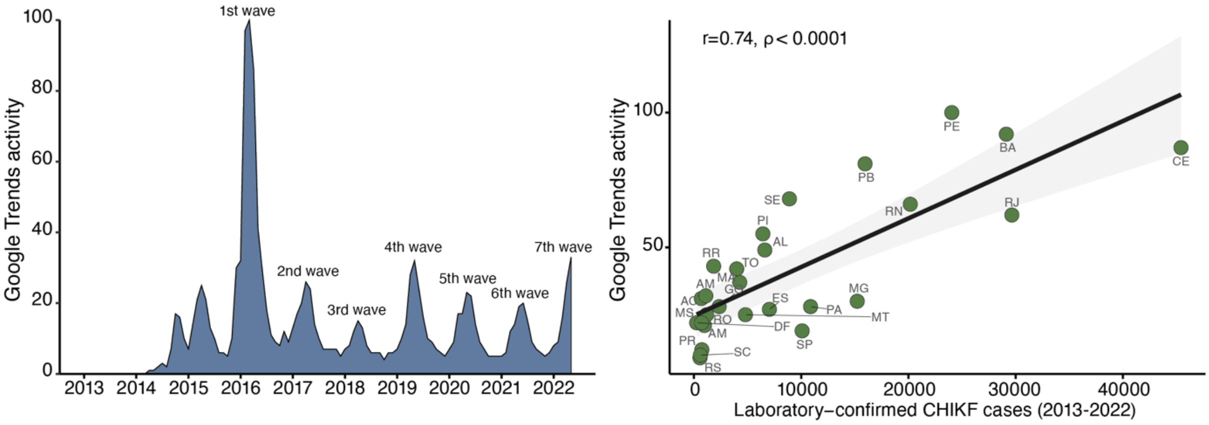


**Fig. S1. Digital surveillance of chikungunya virus in Brazil**. Timeline of Google Trends activity for the term “chikungunya” in Brazil from January 2013 to May 2022 (left). Correlation between Google Trends activity and laboratory confirmed CHIFV cases per state from January 2013 to May 2022 (right). The correlation was calculated using the Spearman’s rank correlation coefficient. AC, Acre; AL, Alagoas; AM, Amazonas; AP, Amapá; BA, Bahia; CE, Ceará; DF, Distrito Federal; ES, Espírito Santo; GO, Goiás; MA, Maranhão; MG, Minas Gerais; MS, Mato Grosso do Sul; MT, Mato Grosso; PA, Pará; PB, Paraíba; PE, Pernambuco; PI, Piauí; PR, Paraná; RJ, Rio de Janeiro; RN, Rio Grande do Norte; RO, Rondônia; RR, Roraima; RS, Rio Grande do Sul; SC, Santa Catarina; SE, Sergipe; SP, São Paulo; TO, Tocantins.

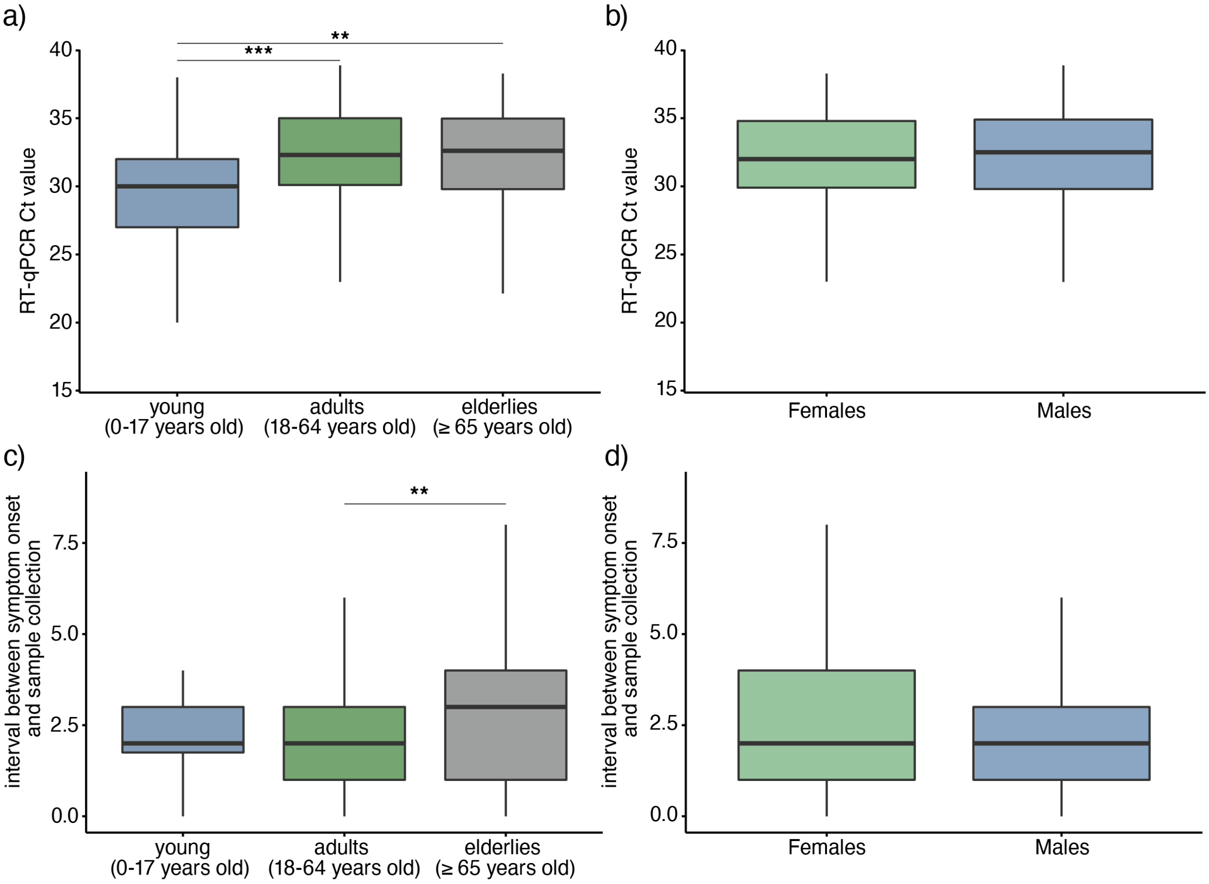


**Fig. S2. Distribution of cycle threshold values and the interval between symptom onset and sample collection at a Laboratory of Public Health of Ceará State.** Cycle threshold values for CHIKF cases were stratified according to the age group (**a**) or sex (**b**). The interval between symptom onset and sample collection per age group (**c**) or sex (**d**). The statistical difference between sexes in all age groups was calculated by One-way ANOVA with the Tukey's HSD (honestly significant difference) test. Significance is indicated as * p < 0.05, ** p < 0.01, *** p < 0.001 or p > 0.05 (ns).

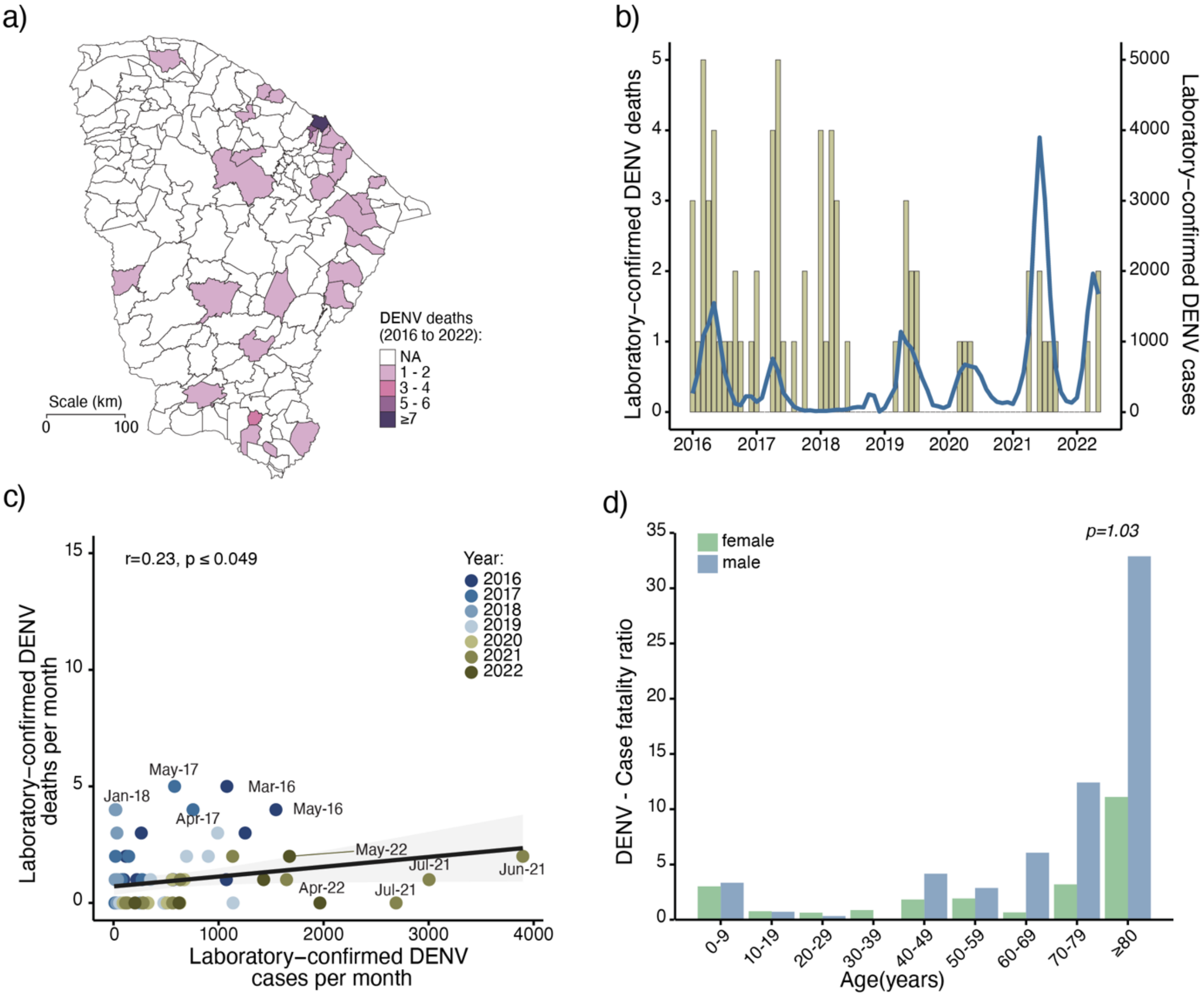


**Fig. S3. Dengue deaths in Ceará State, Brazil. a)** Spatial distribution of laboratory-confirmed DENV deaths per municipality in Ceará State from 2016 to 2022. **b)** Number of laboratory-confirmed DENV deaths (blue line) and cases (gold bars) per month from 1 January 2016 to 31 May 2022. **b)** The Pearson's correlation coefficients of laboratory-confirmed DENV deaths and laboratory-confirmed DENV cases per month from 2016 to 2022 in Ceará State. **d)** The statistical difference between sexes in all age groups was calculated by two-way ANOVA and a post-hoc test were performed correcting for multiple comparisons using the Dunn–Bonferroni method. The significance level was determined as a p-value less than 0.05.

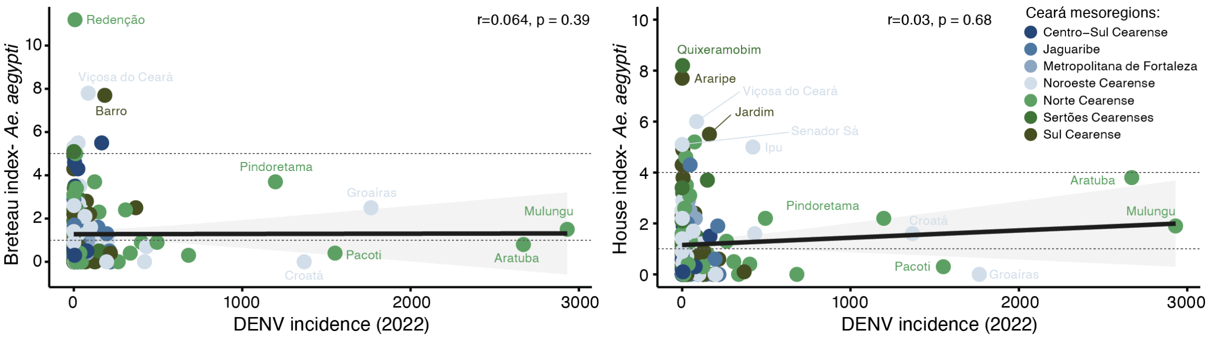


**Fig. S4. Correlation between *Ae. aegypti* population density indices and dengue incidence in Ceará in 2022.** Correlation coefficients of Breteau index (left) and House index (right) per DENV incidence in 2022. The correlation was calculated using Spearman's rank correlation coefficient. The circles were colored based on Ceará mesoregions. The upper dash line indicates the risk situation

(Breteau index = 5 and House index = 4), and the bottom dash line denotes the alert situation (Breteau index and House index > 1) according to the National Dengue Control Programme in Brazil (28).

### Table Supplementary 1. Chikungunya epidemic waves in Brazil.

| Wave | Year | Total cases[1] | States most affected | Number case in wave | Period of CHIFV epidemic peak | Peak of CHIFV cases[3] |
|---|---|---|---|---|---|---|
| 1 | 2016 | 44,604 | Pernambuco | 8,794 (19.7%)[2] | February to March | 539 to 758 |
| | | | Ceará | 11,474 (27.7%)[2] | April to June | 575 to 835 |
| 2 | 2017 | 41,974 | Ceará | 21,486 (51.2%)[2] | March to July | 535 to 1,692 |
| 3 | 2018 | 24,097 | Rio de Janeiro | 6,316 (26.2%)[2] | April | 329 to 392 |
| 4 | 2019 | 33,740 | Rio de Janeiro | 15,984 (47.4%)[2] | April to June | 581 to 1,087 |
| 5 | 2020 | 31,233 | Bahia | 12,390 (39.7%)[2] | May to June | 562 to 620 |
| 6 | 2021 | 39,224 | Pernambuco | 6,628 (16.9%)[2] | June to July | 311 to 407 |
| | | | São Paulo | 6,539 (16.7%)[2] | March to April | 304 to 390 |
| 7 | 2022 | 35,183 | Ceará | 8,413 (23.9%)[2] | March to May | ≥ 500* |

Legend: [1]Total of CHIKF cases reported in Brazil to the Ministry of Health. [2]Percentage of CHIFV cases in the state related to the total cases in the wave. [3] Number of CHIKF cases per epidemiological week in the peak of epidemic in the state. *Ongoing CHIKF epidemic.

### Table Supplementary 2. Information of chikungunya cases sequenced in this study.

| ID | Sample | Sex | Municipality | Onset symptoms | Sample collection | Ct value - RT-PCR |
|---|---|---|---|---|---|---|
| 1 | serum | M | Barbalha | Feb-22 | Feb-22 | 23.3 |
| 3 | serum | F | Barbalha | Feb-22 | Feb-22 | 21.94 |
| 4 | serum | F | Barbalha | Feb-22 | Feb-22 | 29.55 |
| 5 | serum | M | Barbalha | Feb-22 | Feb-22 | 25.14 |
| 6 | serum | F | Barbalha | Feb-22 | Feb-22 | 29.03 |
| 7 | serum | M | Barbalha | Feb-22 | Feb-22 | 23.14 |
| 8 | serum | M | Barbalha | Feb-22 | Feb-22 | 25.36 |
| 9 | serum | M | Barbalha | Feb-22 | Feb-22 | 26.15 |
| 10 | serum | M | Barbalha | Feb-22 | Feb-22 | 25.49 |
| 11 | serum | F | Barbalha | Feb-22 | Feb-22 | 21.63 |
| 12 | serum | F | Barbalha | Feb-22 | Feb-22 | 23.33 |
| 13 | serum | M | Barbalha | Feb-22 | Feb-22 | 22.9 |
| 14 | serum | M | Barbalha | Feb-22 | Feb-22 | 28.96 |
| 15 | serum | F | Barbalha | Feb-22 | Feb-22 | 27.69 |
| 16 | serum | M | Barbalha | Feb-22 | Feb-22 | 22.13 |
| 17 | serum | F | Barbalha | Feb-22 | Feb-22 | 29.35 |
| 18 | serum | M | Barbalha | Feb-22 | Feb-22 | 27.17 |
| 20 | serum | F | Barbalha | Feb-22 | Feb-22 | 28.49 |
| 21 | serum | F | Barbalha | Feb-22 | Feb-22 | 27.73 |
| 22 | serum | F | Barbalha | Feb-22 | Feb-22 | 29.68 |
| 23 | serum | M | Barbalha | Feb-22 | Feb-22 | 26.49 |
| 24 | serum | F | Barbalha | Feb-22 | Feb-22 | 29.27 |
| 25 | serum | F | Fortaleza | Mar-22 | Mar-22 | 29.6 |

| | | | | | | |
|---|---|---|---|---|---|---|
| 45 | serum | M | Fortaleza | Mar-22 | Mar-22 | 28.8 |
| 46 | serum | F | Pacoti | Feb-22 | Mar-22 | 30.82 |
| 47 | serum | F | Pacoti | Mar-22 | Mar-22 | 30.64 |
| 48 | serum | M | Pacoti | Mar-22 | Mar-22 | 30.63 |
| 50 | serum | F | Pacoti | Mar-22 | Mar-22 | 29.13 |
| 52 | cerebrospinal fluid | M | Fortaleza | Feb-22 | Mar-22 | 29.6 |
| 53 | serum | F | Barbalha | Feb-22 | Feb-22 | 24.51 |
| 54 | serum | F | Pacoti | Feb-22 | Feb-22 | 28.68 |
| 55 | serum | M | Barbalha | Feb-22 | Feb-22 | 29.94 |
| 56 | serum | F | Barbalha | Feb-22 | Feb-22 | 29.93 |
| 57 | serum | F | Pacoti | Feb-22 | Feb-22 | 29.59 |
| 58 | serum | M | Barbalha | Feb-22 | Feb-22 | 18 |
| 59 | serum | F | Barbalha | Feb-22 | Feb-22 | 23 |
| 60 | serum | F | Barbalha | Feb-22 | Feb-22 | 20 |
| 61 | serum | M | Barbalha | Feb-22 | Feb-22 | 21 |
| 62 | serum | M | Barbalha | Feb-22 | Feb-22 | 24 |
| 63 | serum | F | Barbalha | Feb-22 | Feb-22 | 27 |
| 64 | serum | M | Barbalha | Feb-22 | Feb-22 | 20 |
| 65 | serum | F | Barbalha | Feb-22 | Feb-22 | 20 |
| 66 | serum | M | Barbalha | Feb-22 | Feb-22 | 17 |
| 67 | serum | F | Barbalha | Feb-22 | Feb-22 | 21 |
| 68 | serum | F | Penaforte | Feb-22 | Feb-22 | 20 |
| 70 | serum | F | Juazeiro do Norte | Mar-22 | Mar-22 | 28 |
| 72 | serum | F | Juazeiro do Norte | Mar-22 | Mar-22 | 18 |
| 73 | serum | F | Farias Brito | Mar-22 | Mar-22 | 25 |
| 74 | serum | F | Juazeiro do Norte | Mar-22 | Mar-22 | 23 |
| 75 | serum | F | Juazeiro do Norte | Mar-22 | Mar-22 | 19 |
| 76 | serum | F | Juazeiro do Norte | Mar-22 | Mar-22 | 27 |
| 77 | serum | F | Juazeiro do Norte | Mar-22 | Mar-22 | 21 |
| 78 | serum | F | Juazeiro do Norte | Mar-22 | Mar-22 | 23 |
| 79 | serum | F | Juazeiro do Norte | Mar-22 | Mar-22 | 21 |
| 80 | serum | M | Juazeiro do Norte | Mar-22 | Mar-22 | 20 |
| 81 | serum | M | Juazeiro do Norte | Mar-22 | Mar-22 | 30 |
| 82 | serum | M | Juazeiro do Norte | Mar-22 | Mar-22 | 25 |
| 83 | serum | M | Juazeiro do Norte | Mar-22 | Mar-22 | 20 |
| 84 | serum | F | Juazeiro do Norte | Mar-22 | Mar-22 | 22 |
| 85 | serum | F | Juazeiro do Norte | Mar-22 | Mar-22 | 21 |
| 86 | serum | M | Juazeiro do Norte | Mar-22 | Mar-22 | 24 |

Legend: F, female. M, male.
